# A randomized, controlled, feasibility study of RD-X19 in patients with mild-to-moderate COVID-19 in the outpatient setting

**DOI:** 10.1101/2021.10.17.21265058

**Authors:** Nathan Stasko, Adam S. Cockrell, Jacob F. Kocher, Ibrahim Henson, David Emerson, Ye Wang, Jonathan R. Smith, Nathan H. Henderson, Hillary Wood, Shelton S. Bradrick, Terry Jones, Jorge Santander, John G. McNeil

## Abstract

These studies aimed to further understand the antiviral effects of safe, visible light and demonstrate a therapeutic effect of an investigational treatment device for outpatients with mild to moderate COVID-19. RD-X19 is a handheld medical device precisely engineered to emit blue light through the oral cavity to target the oropharynx and surrounding tissues. At doses that are well-tolerated in an *in vitro* human epithelial tissue model, the monochromatic visible light delivered by RD-X19 results in light-initiated expression of IL-1α and IL-1β cytokines with corresponding inhibition of SARS-CoV-2 replication. A randomized, double-blind, sham-controlled early feasibility study using the investigational device enrolled 31 subjects with a positive SARS-CoV-2 antigen test and possessing at least two moderate COVID-19 signs and symptoms. Subjects were randomized 2:1 (RD-X19 to sham), treated twice daily for four days, and evaluated over one week. Prespecified outcome measures included assessments of SARS-CoV-2 viral load and clinical assessments of COVID-19. There were no local application site reactions and no device-related adverse events. The time-weighted average change in log viral load throughout the study demonstrated a favorable reduction for RD-X19 compared to sham and at the end of study the mean change in log_10_ viral load was -3.29 for RD-X19 and -1.81 for sham at Day 8, demonstrating a treatment benefit of -1.48 [95% confidence internal (CI), -2.88 to -0.071]. Among the clinical outcome measures, differences between RD-X19 and sham were also observed, with a 57-hour reduction of median time to sustained resolution of COVID-19 signs and symptoms.

## INTRODUCTION

Severe acute respiratory syndrome coronavirus type 2 (SARS-CoV-2) genetic variants continue to propagate the global COVID-19 pandemic. As diagnostic procedures have evolved throughout the pandemic, reports have demonstrated that SARS-CoV-2 viral load measured in saliva strongly correlates with disease severity and mortality.^1-3^ Correspondingly, the importance of the oral cavity in both disease progression and oral-lung transmission via aspiration is now clear, with many tissues and glands in the oral cavity having high levels of ACE2 and TMPRSS2 expression required for SARS-CoV-2 entry into barrier epithelia.^4-5^

Systemically administered antibody cocktails and convalescent plasma have shown clinical evidence of viral load reductions and improved clinical outcomes in non-hospitalized populations;^6-8^ however, they are indicated for populations with risk factors for progression to severe disease/hospitalization and require infusion in a clinical setting, limiting these treatments to roughly 25% of the infected population.^9^ Further, novel variants of concern such as the currently circulating Delta variant will continue to emerge with potential enhancements in transmissibility and resistance to existing antibody therapies and vaccines,^10-12^ underscoring the need for innovative therapeutics directly targeting the virus in the nasal or oral cavity early in the disease process.

Light therapy has successfully been used for many years as a treatment for skin disorders^13^ but has yet to be successfully adapted to respiratory medicine. Several potential mechanisms of action have been postulated following host-directed photobiomodulation including the release of endogenous nitric oxide^14^ and alteration of cellular redox states that activate transcription factors and other immune signaling pathways.^15^ In the exploration of light as an antiviral, Zupin et. al. administered 10-20 J/cm^2^ doses of various blue light wavelengths (450, 454, and 470 nm) that were non-toxic to cells *in vitro* and reduced SARS-CoV-2 viral replication for up to 48 hours post infection.^16^ Effects for all three wavelengths were observed once the virus entered the cells, suggesting that blue light may interfere with the intracellular viral replication. Recently, Cockrell and co-workers reported that 425 nm blue light dramatically inhibited SARS-CoV-2 infection and replication in primary human 3D tracheal/bronchial tissue at doses (≤ 32 J/cm^2^) that are well tolerated by epithelial tissues.^17^

Safe, non-UV light therapy is neither antigen-directed nor antigen-dependent, affording a novel therapeutic approach that can be widely deployed to mitigate the threat posed by current SARS-CoV-2 variants, pre-emergent coronaviruses, and potentially other non-coronavirus pathogens. Herein we report confirmatory evidence of the antiviral effects of 425 nm blue light in SARS-CoV-2 infected oral epithelial tissues and translation of this technology into an investigational device, RD-X19, where a twice-daily dosing schedule was evaluated in a randomized, controlled early feasibility study in outpatients with mild to moderate COVID-19.

## METHODS

### Oral Epithelial Tissue Model

Primary oral mucosal tissues derived from human buccal epithelial cells (EpiOral™ ORL-200, MatTek Corporation – 40 year old Caucasian male tissue donor) were cultured for 28 days in transwell inserts as described previously.^18^ Following transport, upon arrival, the transwell inserts were removed and placed in hanging well plates with ORL-200-MM maintenance media in the basal compartment; no media added to the apical surface. Tissues were incubated at 37°C and 5% CO_2_ overnight prior to experimental use.

#### SARS-COV-2 Infected Tissues

For SARS-CoV-2 infections of ORL-200 tissue cultures, 200 μL of SARS-CoV-2 WA1/2020 P4 diluted in virus diluent at an MOI of 0.1 [MEM supplemented with 2% FBS (Gibco), 1% nonessential amino acids (Gibco), and 1% antibiotic-antimycotic (Gibco)] was inoculated on the apical surface and incubated for 2 hours at 37°C and 5% CO_2_. Virus was then removed from the apical surface and the transwell cultures were further incubated for 1 additional hour at 37°C and 5% CO_2_ until administration of the first dose of light. Infected transwell cultures were illuminated at room temperature with 425 nm light at doses of 0 J/cm^2^, 16 J/cm^2^, 32 J/cm^2^, or 60 J/cm^2^ using LED array biological light units and viral titers were measured from apical washes taken 12 and 24 hours post infection as described previously.^17^ All work with live virus was conducted in a CDC certified, BSL-3 laboratory at EmitBio with adherence to established safety guidelines.

#### Tissue Viability

Using separate, un-infected transwell cultures, the cytotoxicity of each dose of light was assayed at 24 hours post exposure using 3-(4,5-dimethylthiazol-2-yl)-2,5-diphenyltetrazolium bromide (MTT) following manufacturer’s instructions (MTT-100, MatTek corporation). Briefly, tissues were rinsed with TEER buffer and placed into 300 μL of pre-warmed MTT reagent and incubated at 37 °C and 5% CO_2_ for 3 hours. The MTT stained tissue inserts were then extracted for 2 hours and the absorbance of 200 μL of extract solution was measured at 560 nm on a GloMax Discover (Promega). The viability of light-exposed tissues was calculated relative to dark controls using the following equation: Relative viability = [OD_sample_/Mean OD_neg_ ctrl] x 100.

#### Gene Expression Analysis

To assess the immunomodulatory effects of 425 nm light on oral epithelial tissues, un-infected ORL-200 transwell cultures were illuminated with a single dose of 425 nm light (0 J/cm^2^, 3 J/cm^2^, 7.5 J/cm^2^, 15 J/cm^2^, 30 J/cm^2^, or 60 J/cm^2^) and incubated at 37°C, 5% CO_2_. After 24 hours, ORL-200 tissues were lysed with the QuantiGene Sample Processing Kit for Cultured Cells (Thermo Scientific) following manufacturer’s instructions for “Cells grown in 3D cell matrix.” Lysates were stored individually at -80 °C until hybridization and RNA expression analysis on the Magpix® Instrument System (Luminex Corporation) according to manufacturer instructions. Gene expression was normalized to the geometric mean of 3 housekeeping genes (PPIB, TBP, and HPRT1) and presented as fold change over mock-illuminated tissues (0 J/cm^2^) +/-SD.

### Early Feasibility Clinical Study Design

This randomized, double-blind, sham-controlled, early feasibility study was conducted in the outpatient setting at two centers in the United States. Subjects were required to have symptomatic COVID-19 and active SARS-CoV-2 infection confirmed by an FDA-approved rapid antigen test. The trial was conducted in accordance with the principles of the Declaration of Helsinki, International Council for Harmonization of Good Clinical Practice guidelines and all applicable regulatory requirements including oversight by a local institutional review board for each trial center. All subjects provided written informed consent before participating in the trial. ClinicalTrials.gov Identifier: NCT04662671

#### Subjects

Eligible subjects were 18 to 65 years old with positive results on testing for SARS-CoV-2 via an FDA-authorized rapid antigen test within 24 hours of enrollment. Entry criteria also required participants to have either a fever of at least 100°F or at least two of the eight protocol-designated signs and symptoms of COVID-19 graded as moderate or higher. Signs and symptoms were cough, sore throat, nasal congestion, headache, unexplained chills or sweats, muscle or joint pain, fatigue, and nausea where each was assessed on a 4-point scale (symptom score of 0=absent, 1=mild, 2=moderate, and 3=severe). The Composite Severity Score (ranging from 0 to 3 and ≥0.5 at baseline) was defined as the sum of the eight individual COVID-19 signs and symptoms severity scores divided by eight. Individuals displaying these signs and symptoms longer than three days, with BMI ≥36 kg/m^2^, or with COVID-19 signs or symptoms indicative of imminent acute respiratory distress syndrome or severe COVID-19 were excluded. A full listing of inclusion and exclusion criteria can be found in Supplementary Information.

#### Randomization and Intervention

All participants were centrally randomized using randomly permuted blocks of six. The subjects, investigators, site personnel, and EmitBio employees who were involved in collecting and analyzing data were unaware of the treatment-group assignments. A total of 31 study subjects were randomized 2:1 (RD-X19:Sham). Each study subject was trained how to insert the device into the mouth and instructed to treat twice daily for a duration of four days. The RD-X19 treatment times were controlled via a pre-programmed timer integrated into the RD-X19 device to deliver a nominally designed light dose of 16 J/cm^2^ per treatment to the oropharynx. The sham devices were designed to provide the same user experience but at energy and fluence levels with a lower inactivation potential against SARS-CoV-2 *in vitro* than that delivered by the active devices. Illumination throughout the oral cavity and the nominal light doses provided by RD-X19 as determined by optical modeling are illustrated in Figure 1 (Methods S1). The nominal RD-X19 light dose of 16 J/cm^2^ and BID treatment schedule was selected based on *in vitro* evidence generated using SARS-CoV-2 infected large airway epithelial tissues^17^ and supporting safety data generated in a prior Phase 1 trial of RD-X19 with similar total doses of energy on a weekly basis (NCT04557826).

**Figure 1.**
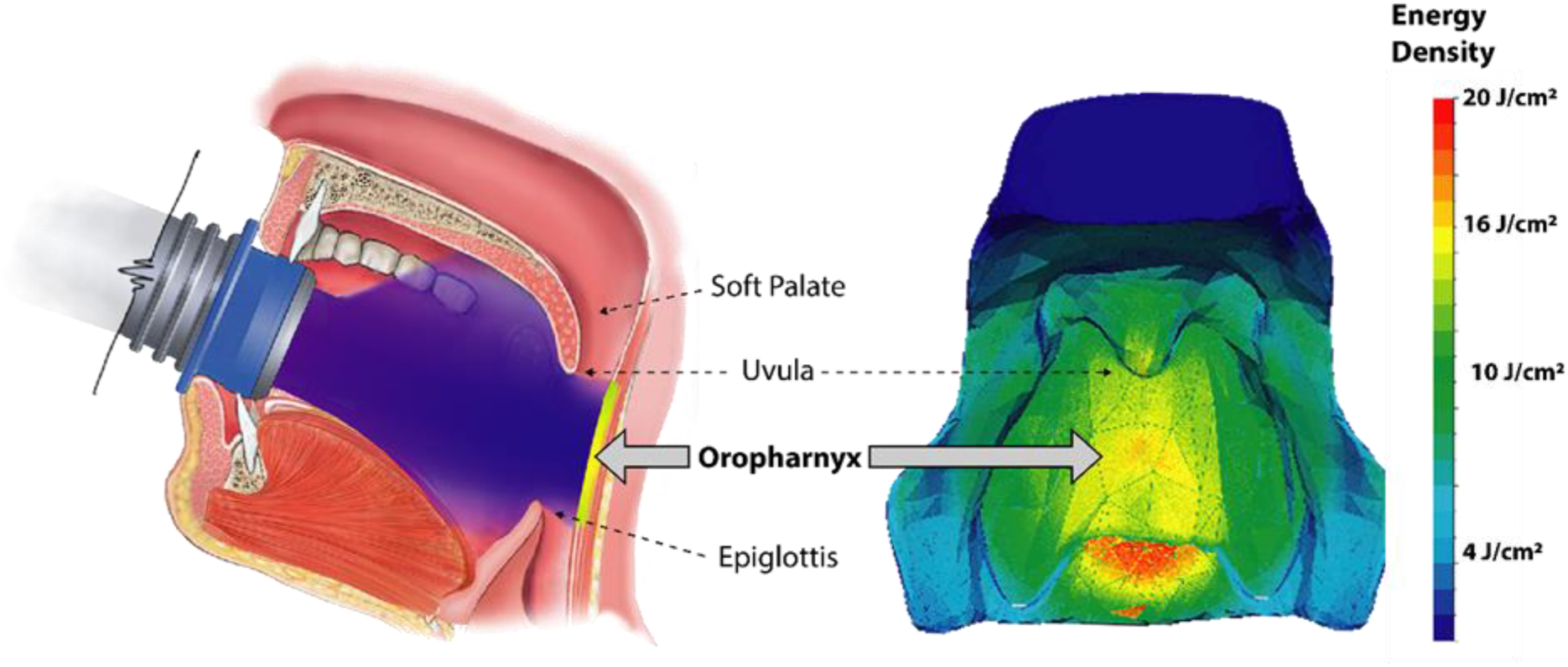
RD-X19 Intervention and Optical Modeling of Energy Density in the Oral Cavity. A) Sagittal view showing a schematic RD-X19 device positioned in the oral cavity to deliver a dose of light to the oropharynx and surrounding tissues. The blue shading is included as a guide to the eye to illustrate the approximate shape of the light beam and coverage of anatomical features. B) Results of optical modeling of nominally designed light dose of 16 J/cm^2^ emitted from a typical RD-X19 device and projected onto the surfaces of the oral cavity. Elements of the anatomical model (e.g. cheeks) have been removed for better visualization of the light projection. LightTools, a Monte-Carlo-based ray-tracing program for 3D simulations of complex systems with the ability to incorporate volumetric optical effects, scattering, and absorption throughout the oral cavity, was used to generate the dose contour plot (Methods S1).

### Safety Assessments

Safety assessments included evaluation of vital signs, local applications site reactions via oropharyngeal exam, treatment emergent adverse events (TEAEs), and serious adverse events (SAEs). For safety and tolerability, local site reactions (pain, induration, erythema) and the presence, type, severity, and attribution of any device-related TEAEs were assessed. Hematological, metabolic, liver, and kidney function evaluations were also performed at baseline and at day 8.

### Outcome Assessments

#### Virologic Outcomes

The primary outcome was the time-weighted average change (TWAC) in SARS-CoV-2 viral load from baseline through day 8 as measured by saliva specimens obtained from each participant via OMNIgene Oral collections kits (OM-505, DNA Genotek) and analyzed by quantitative reverse-transcriptase-polymerase-chain-reaction (RT-qPCR) testing at a central laboratory (log_10_ copies per milliliter, Methods S2). Other prespecified key virologic outcome measures included the change in viral load from baseline (D1) at days 3, 5, and 8 (D3, D5, D8) and proportion of subjects with >95% clearance or undetectable levels of SARS-CoV-2 in saliva.

#### Clinical Outcomes

Clinical outcomes included time to event analyses of symptoms resolution and change in Composite Severity Score from baseline. Participants self-assessed severity of the eight protocol-designated COVID-19 signs and symptoms twice daily via diary cards (Figure S1). The time (measured in hours) to alleviation of COVID-19 signs and symptoms was defined as the point at which the eight COVID-19 signs and symptoms scores were first reported by the participant to all be 0 (none) or 1 (mild). The time to sustained resolution of COVID-19 was established as a post hoc analysis, incorporated FDA Guidance,^17^ where *sustained* symptom resolution uses the same ‘success’ definition above but *without rebound of any score >1 for the remainder of the trial*.

### Sample Size and Statistical Analyses

This was a hypothesis generating, proof-of-concept trial where the number of participants was chosen to provide a preliminary assessment of safety and efficacy. All clinical and safety analyses were conducted on the intent-to-treat (ITT) population. All virologic efficacy assessments were conducted on a modified analysis set including all study subjects with laboratory confirmed SARS-CoV-2 positivity via RT-qPCR at baseline. The prespecified primary efficacy endpoint of TWAC in log_10_-transformed viral load was derived using the trapezoidal rule and the RD-X19 compared to sham using an Analysis of Covariance (ANCOVA) model with baseline viral load on log_10_ scale as a covariate and treatment group as an independent variable using two-sided tests with an alpha level of 0.05. Kaplan-Meier product limit estimates to compare the time to reach symptom resolution endpoints were performed using log-rank tests. No adjustments for multiplicity were conducted for this study. Statistical analyses were performed with SAS software, version 9.4 (SAS Institute Inc.).

## RESULTS

### Photobiomodulation of Oral Mucosal Tissue with 425 nm Light

Based on the evidence generated previously on SARS-CoV-2 infected tracheal/bronchial airway epithelial tissues,^17^ experiments were conducted to evaluate whether SARS-COV-2 replicates in oral epithelia and to evaluate whether doses of 425 nm light could inhibit virus replication with minimal cellular toxicity. Similar to data reported using primary human large airway tissues, single exposures of 16, 32, or 60 J/cm^2^ of 425 nm light all significantly inhibited infection and replication of the parental SARS-CoV-2 (WA1) in oral epithelial tissues at 12- and 24-hours post-infection (Fig. 2A). Only the lowest dose tested, 16 J/cm^2^ had any detectable virus at both timepoints evaluated. The minimum dose was chosen based on results from a Phase 1 trial which demonstrated that a nominal total daily dose of 18 J/cm^2^ could be safely self-administered in an at-home setting for up to 2 weeks in duration (NCT04557826). Commensurate with the previously observed tolerability data, single exposures up to 60 J/cm^2^ demonstrated no loss in tissue viability after 24 hours (Figure 2B).

**Figure 2.**
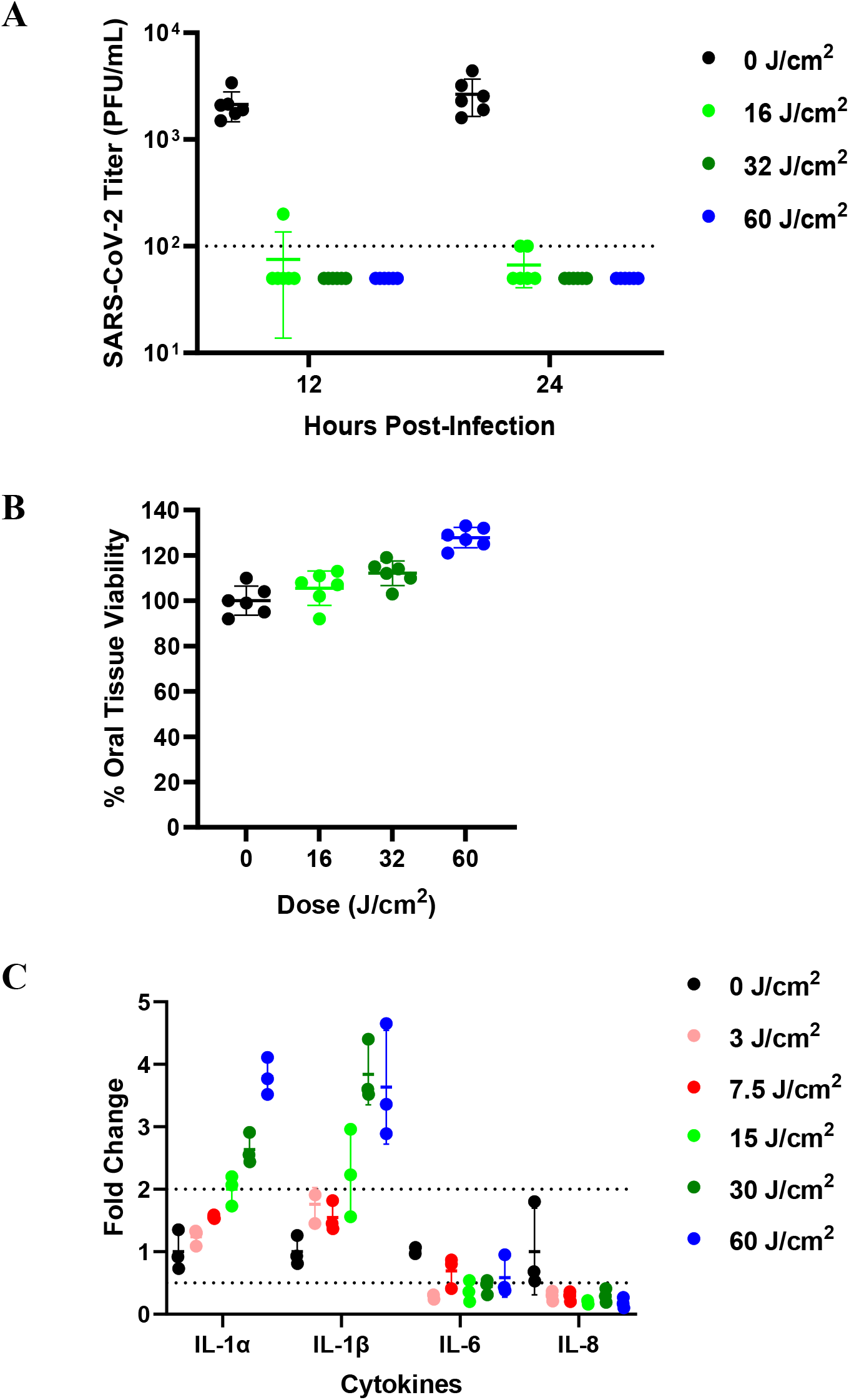
Safe doses of 425nm light inhibits SARS-CoV-2 replication in ORL-200 tissues and induce host immune responses. A) Reduction in SARS-CoV-2 Viral Titer assessed at 12 and 24 hours following a single exposure of 425 nm blue light at doses of 16 J/cm^2^ (light green), 32 J/cm^2^ (dark green), and 60 J/cm^2^ (blue) compared to untreated infected ORL-200 tissue cultures (0 J/cm^2^). Mean ±SD, n=6 independent tissue replicates for each assessment. B) Tissue Viability of uninfected ORL-200 cultures assessed at 24 hours post exposure to light. Mean ±SD, n=6 independent tissue replicates for each assessment. C) Dose dependent fold change in cytokine RNA expression over mock illuminated tissues (0 J/cm^2^) 24 hours after light exposure for a range of 425 nm blue light doses. Mean ±SD, n=3 independent tissue replicates for each assessment.

Gene expression analysis was also conducted on uninfected ORL-200 epithelial tissues illuminated with doses ranging from 3 – 60 J/cm^2^ to assess biomarkers of cellular stress and key host immune response cytokines. As show in Figure 2C, dose dependent increases in Interleukin-1 cytokines (IL-1α and IL-1β) were observed with >3-fold increase after a single illumination of 60 J/cm^2^. Blue light appears to have a selective impact on certain promoters of innate immune response with no corresponding increases of IL-6 or IL-8. Importantly, no dose dependent increases were observed in several markers of cellular (CASP3, LDH-B), metabolic (HSPD1), and oxidative stress (NRF2, KEAP1) with the same dosing schedule of light (Figure S2A).

### Demographics and Baseline Disease Burden

Subjects were randomized within three days or less from the onset of COVID-19 signs and symptoms. Demographics, baseline disease characteristics, and baseline saliva viral load are listed in Table 1. Of the 31 study participants who underwent randomization, 20 received RD-X19 and 11 received the sham device (Figure 3). The subjects who tested positive for SARS-CoV-2 in saliva by RT-qPCR at randomization (28 of 31, 90%) comprised the modified analysis set for virologic endpoint analyses. The mean baseline Composite Severity Score was 1.29 (SD, 0.35) for the total study population and was similarly balanced between treatment groups.

**Table 1.** Demographic and Baseline Medical Characteristics.

| Characteristic | Category | RD-X19<br>Treatment<br>(N=20) | Sham<br>Device<br>(N=11) | Total<br>(N=31) |
| --- | --- | --- | --- | --- |
| **Demographics** |  |  |  |  |
| Age | Median | 43 | 36 | 40 |
|  | Min, Max | 20, 65 | 21, 57 | 20,65 |
| Gender, no. (%) | Male | 8 (40%) | 8 (73%) | 16 (52%) |
|  | Female | 12 (60%) | 3 (27%) | 15 (48%) |
| Race or ethnic group, no. (%)<sup>a</sup> | Hispanic or Latino | 15 (75%) | 7 (64%) | 22 (71%) |
|  | White | 4 (20%) | 3 (27%) | 7 (23%) |
|  | Black | 1 (5%) | 1 (9%) | 2 (6%) |
| Body Mass Index (kg/m<sup>2</sup>) | Median | 28.8 | 29.1 | 28.8 |
|  | Min, Max | 22.3, 35.1 | 22.1, 35.4 | 22.1, 35.4 |
| **Disease Characteristics** |  |  |  |  |
| Risk Factors for severe COVID-19, n (%)<sup>b</sup> | Confirmed | 12 (60%) | 4 (36%) | 16 (52%) |
| Composite Severity Score @ BL<sup>c</sup> | Mean | 1.26 | 1.36 | 1.29 |
|  | (SD) | (0.39) | (0.28) | (0.35) |
| Baseline Disease Severity<sup>d</sup> | Mild | 14 (70%) | 8 (73%) | 22 (71%) |
|  | Moderate | 6 (30%) | 3 (27%) | 9 (29%) |
| **Baseline Viral Load in saliva specimen** |  |  |  |  |
| Positive at baseline via RT-qPCR | – no.<br>(%) | 17/20<br>(85%) | 11/11<br>(100%) | 28/31<br>(90%) |
| SARS-CoV-2 cycle threshold<sup>e</sup> | Mean C<sub>t</sub><br>(SD) | 28.9<br>(5.1) | 30.4<br>(4.5) | 29.5<br>(4.8) |
| Mean SARS-CoV-2 Viral Load | Log<sub>10</sub> copies/mL<br>(SD) | 4.10<br>(2.27) | 4.41<br>(1.37) | 4.2<br>(1.5) |
| Median SARS-CoV-2 Viral Load | Log<sub>10</sub> copies/mL<br>(range) | 4.29<br>(0.00 – 8.18) | 4.61<br>(2.18 – 6.36) | 4.3<br>(0.00 – 8.18) |

SARS-CoV-2 indicates severe acute respiratory syndrome coronavirus 2
- Race and ethnicity are not mutually-exclusive and both are documented for subjects who self-identify as such. - CDC risk factors of severe disease included medical history of diabetes, chronic kidney or liver disease, cardiovascular disease, chronic respiratory disease, or immunosuppressive disease. - Based on the 8 symptom domains (cough, sore throat, nasal congestion, headache, unexplained chills or sweats, muscle pain or joint pain, fatigue, and nausea) that were rated from absent (0) to severe (3), which were added together and divided by 8 for an overall composite score (range 0 – 3; symptom score did not include loss of taste or smell) - Definition for ‘moderate’ based on clinical signs suggestive of moderate illness with a corresponding respiratory rate $\geq 20$ /minute and/or a heart rate $\geq 90$ /minute.<sup>19</sup> - The cycle threshold is the number of polymerase chain reaction cycles required for a viral sample to be detected. Lower numbers indicate higher viral RNA in the sample and an increased burden of disease. Values range between 0 and 40.

**Figure 3.**
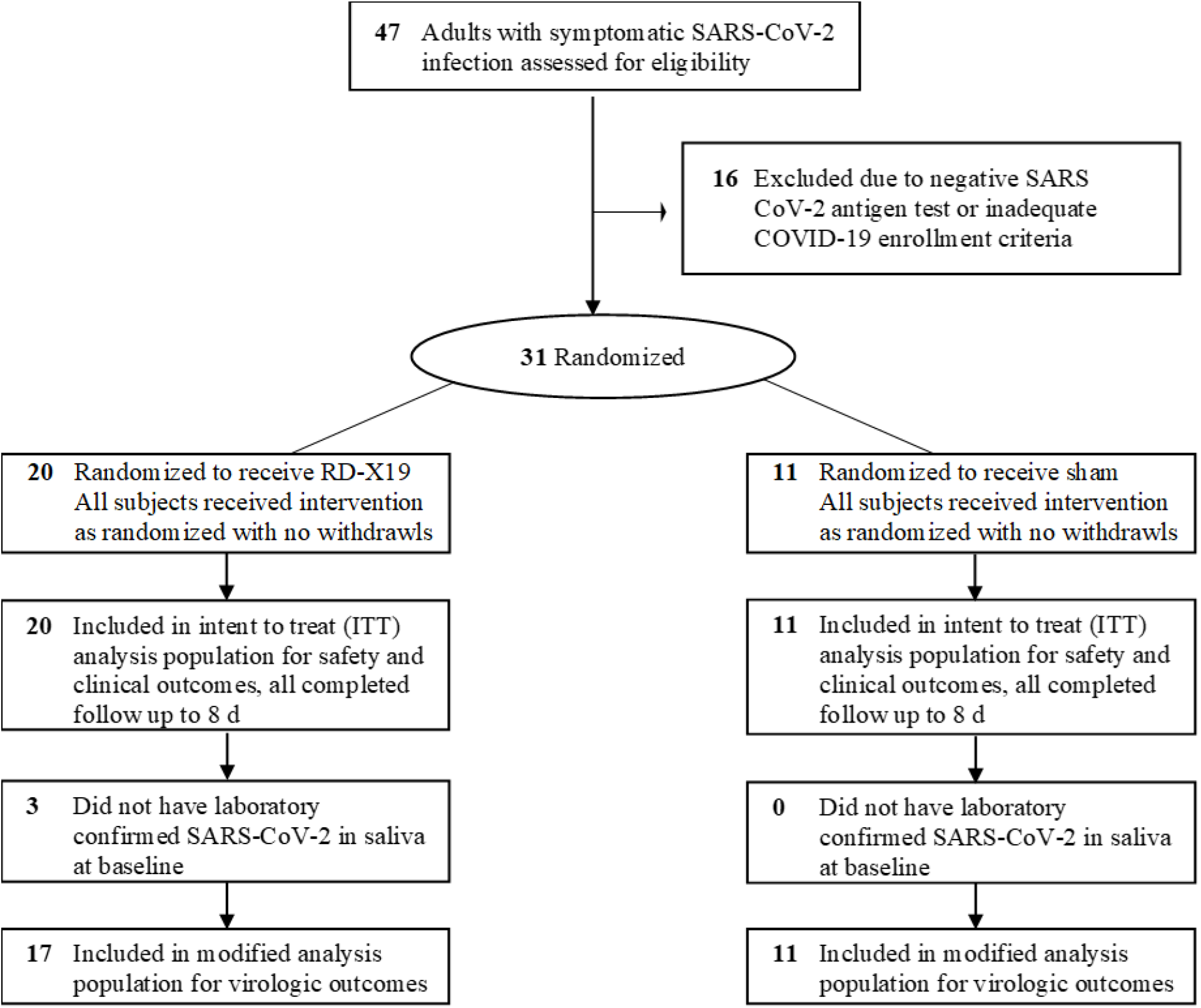
Enrollment and Treatment Assignment. CONSORT Flow diagram illustrating the difference between the intent to treat (ITT, n=31) and the modified analysis population (n=28) with laboratory confirmed SARS-CoV-2 in saliva at baseline.

### Safety and Tolerability

Safety and tolerability were assessed actively at each clinic visit through assessment of vital signs, targeted physical examination, oropharyngeal examination as needed, review of potential TEAEs, and review of daily diary cards. There were no changes in vital signs nor observed adverse oropharyngeal or oral mucosal reactions in any study subject throughout the course of the trial. The device was extremely well tolerated and there were no clinical observations indicative of device-related TEAEs. The most frequently reported adverse events were attributed to perturbations of COVID-19 disease severity, with no discernable difference between treatment groups (Table S1). There were no SAEs and no laboratory values that were out of range with laboratory standards resulting from treatment and no hospitalizations or requirement for acute medical intervention.

### Virologic Outcomes

The prespecified primary outcome measure was TWAC in saliva viral load from baseline through day 8 (D8) where the RD-X19 least-squares mean difference was -0.48 log_10_ copies/mL lower than sham [95% CI, -1.41 to 0.45]. The mean change from baseline (log_10_ transformed data) at each visit for both treatment arms is shown in Figure 4A. On D8, the change in log_10_ viral load for the RD-X19 treated group was -3.29 and for the sham treated group was -1.81, resulting in an RD-X19 group advantage of -1.48 [95% CI, -2.88 to -0.07]. The quantitative determination of SARS-CoV-2 copies/ml determined via RT-qPCR using an RNA standard curve for each individual subject have been provided to illustrate the distribution of SARS-CoV-2 viral load over time (Figure S3). Raw C_t_ values were also normalized to the human RNAse P control gene to calculate mean ΔCt values at baseline and Day 8 for each group as depicted in Figure 4B. The mean change observed in ΔCt for RD-X19 of -7.6 [95% CI, -10.3, -4.9] compared to -4.1 [95% CI, -6.0, -2.2] for sham further illustrates the large reductions in SARS-CoV-2 viral load observed following the RD-X19 treatment regimen.

**Figure 4.**
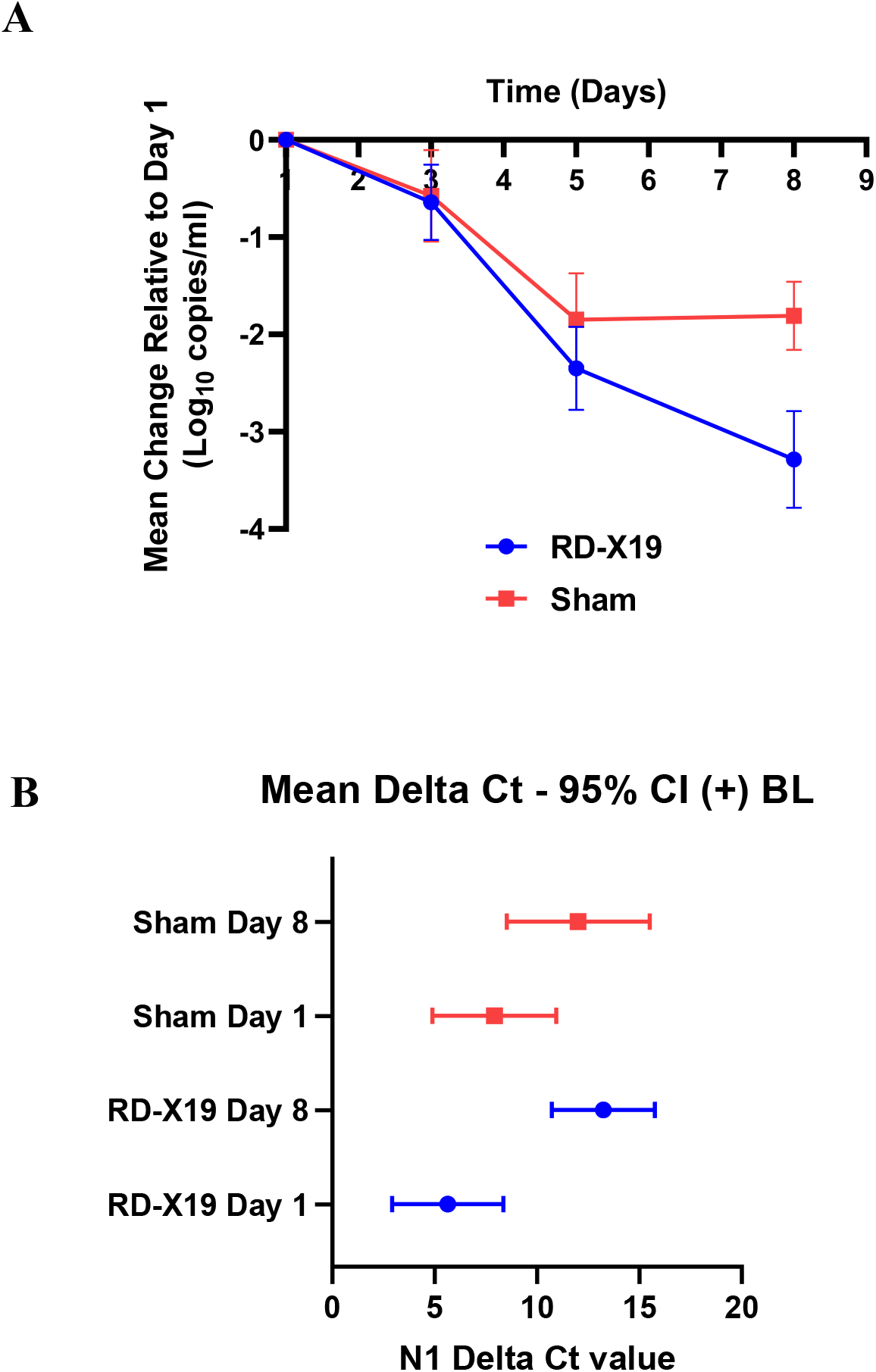
Virologic Outcome Assessments with RD-X19 Therapy. A) Mean change in SARS-CoV-2 saliva viral load (N1 copies/mL) from baseline (Day 1) for subjects receiving treatment with RD-X19 (blue) and subjects receiving sham (red), errors represent +/-SEM. B) Mean change in ΔCt (N1 Ct – RNase P Ct) from baseline for subjects receiving treatment with RD-X19 (blue) versus sham (red).

### Clinical Outcomes

Various clinical outcome assessments were planned to explore the impact of RD-X19 treatment on clinical resolution of COVID-19 signs and symptoms. Key prespecified secondary endpoints were 1) change in Composite Severity Score from baseline and 2) median time to alleviation of symptoms. On average, subjects treated with RD-X19 experienced a greater reduction of the Composite Severity Score at each assessment during the trial (Table 2) with a larger proportion of subjects achieving sustained resolution of symptoms by day 8.

**Table 2.** Summary of Key Virologic and Clinical Outcomes.

|  | **RD-X19** | **SHAM** |
| --- | --- | --- |
| **VIROLOGIC OUTCOMES** | **N=17** | **N=11** |
| **Time-Weighted Average Change (N1 Log<sub>10</sub> copies/mL)** |  |  |
| Least squares mean | -1.69 | -1.23 |
| 95% CI | (-2.27, -1.12) | (-1.94, -0.52) |
| Difference vs. Sham at day 8 | -0.47 | -- |
| 95% CI | (-1.39, 0.45) |  |
| **Saliva Viral Load Over Time (N1 Log<sub>10</sub> copies/mL)** |  |  |
| Mean change from baseline at day 3 | -0.64 | -0.57 |
| Mean change from baseline at day 5 | -2.35 | -1.85 |
| Mean change from baseline at day 8 | -3.29 | -1.81 |
| Difference vs. Sham at day 8 | -1.48 | -- |
| **Median SARS-CoV-2 Viral Load on Day 8 (Log<sub>10</sub>)** | 0.00 | 3.17 |
| **Proportion of subjects achieving ≥95% reduction in viral load** | 50% | 36% |
| **CLINICAL OUTCOMES** | **N=20** | **N=11** |
| **Composite Severity Score** |  |  |
| Mean change from baseline at day 3 | -0.63 | -0.44 |
| Mean change from baseline at day 5 | -0.82 | -0.75 |
| Mean change from baseline at day 8 | -1.04 | -0.96 |
| **Kaplan Meier Time to Event Symptom Analyses** |  |  |
| Median time to Alleviation - All None (0) or Mild (1) | 75.3 hours | 112.7 hours |
| 95% CI | (48.3, 117.2) | (38.0, 166.2) |
| Proportion of Subjects Achieving Success | 85% | 82% |
| Median time to Sustained Resolution - All None (0) or Mild (1) without rebound of any score >1 for the remainder of the trial | 103.8 hours | 160.7 hours |
| 95% CI | (69.0, 130.8) | (38.0, NE) |
| Proportion of Subjects achieving success | 85% | 55% |

The Kaplan-Meier analysis for median time to symptom alleviation resulted in a median time to success for RD-X19 of 75.3 hours [95% CI, 48.3 to 117.2] compared with 112.7 hours [95% CI, 38.0 to 166.2] for the sham treatment group (Figure 5A). This corresponds to RD-X19 yielding a 37-hour decrease in median time to symptom resolution compared with sham. Data from a posthoc analysis conducted on study subjects achieving *sustained* resolution of symptoms is shown for both treatment groups in Figure 5B. At the end of the trial, 17 of 20 subjects (85%) in the RD-X19 group had achieved sustained resolution, compared with 6 of 11 subjects (55%) in the sham group. From the Kaplan-Meier analyses, the median time to sustained resolution was 103.8 hours [95% CI, 69.0 to 130.8] for RD-X19 compared with 160.7 hours [95% CI, 38.0 to NE] for sham. This corresponds to a clinically meaningful 57-hour decrease in median time to sustained resolution for RD-X19 compared to sham.

**Figure 5.**
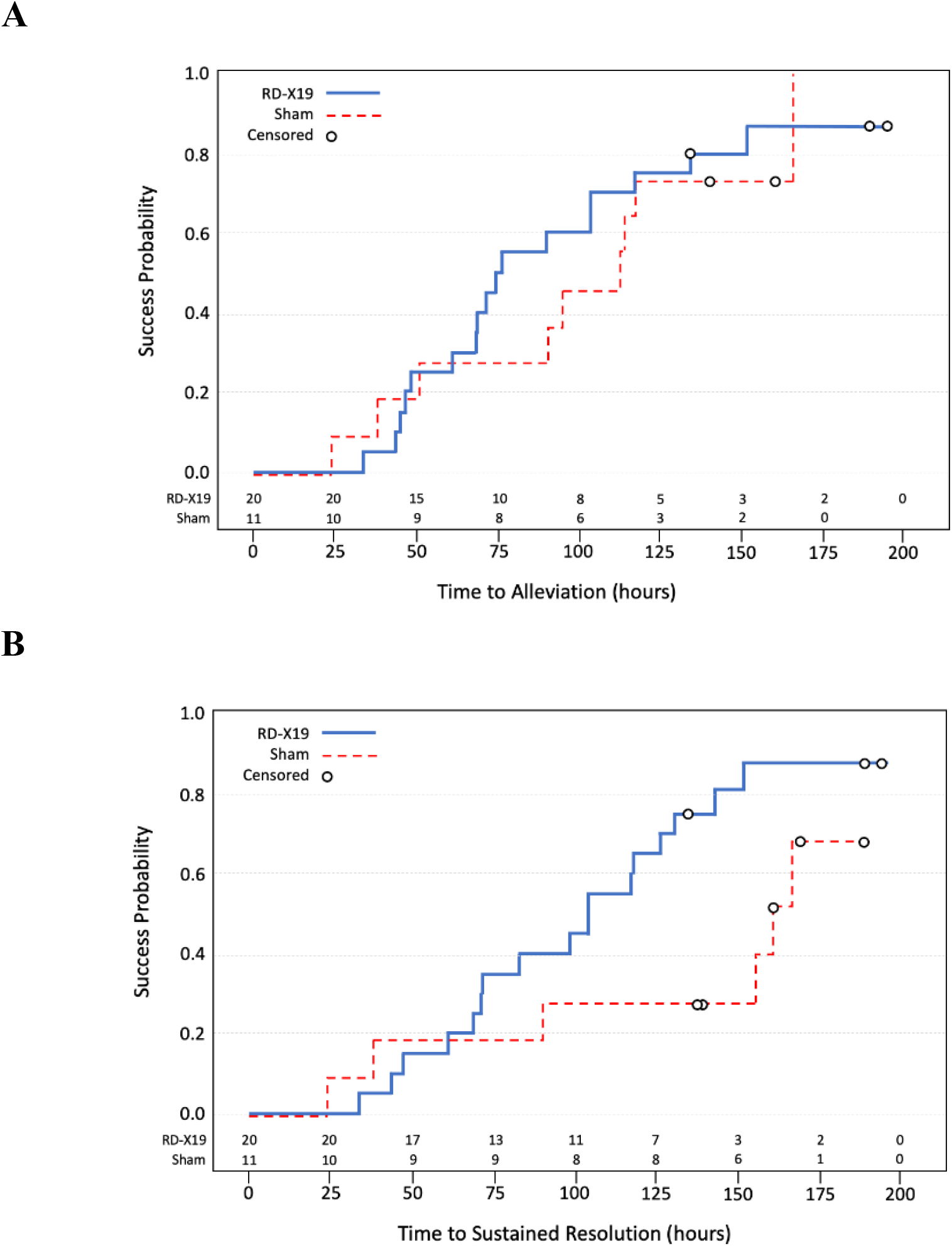
Kaplan Meier Time to Event Analyses of COVID-19 Symptoms. A) Kaplan Meier time to alleviation of symptom analysis with success defined as the first instance subjects achieved symptom scores of all none (0) or mild (1) post baseline. Median time to resolution of symptoms was 75.3 and 112.7 for RD-X19 and sham treatment arms respectively; a difference of approximately 1.5 days faster for RD-X19. B) Kaplan Meier time to sustained resolution of symptoms analysis with success defined as the first instance subjects achieved symptom scores of all none (0) or mild (1) post baseline, *without rebound of any score >1 for the remainder of the trial*. Median time to resolution of symptoms was 103.8 and 160.7 for RD-X19 and sham treatment arms respectively; a difference of more than 2 days faster for RD-X19.

## DISCUSSION

The oral cavity was originally thought to be a passive conduit for the transmission of SARS-CoV-2 to the lower respiratory tract, but recent findings report multiples lines of evidence for a parallel oral axis in both SARS-CoV-2 infection and transmission.^5,20^ Saliva viral load is not only an indicator of local viral shedding in the oral cavity, but it is also a biomarker of total viral bioburden in the respiratory tract and has been shown to be a predictor of COVID-19 disease outcomes.^1-3^ As an example, Fajnzylber et al. reported that 1 to 2 logs higher initial oropharyngeal viral load upon hospital admission was associated with increased mortality.^21^ It is hypothesized that 1) reductions in saliva viral load coincide with resolution of systemic disease, and 2) that a localized anti-viral light treatment targeted to the oropharynx and surrounding tissues may be effective as treatment for COVID-19 in the outpatient setting.

The orally administered RD-X19 device depicted in Figure 1 was devised to exert both direct effects on viral pathogens in the oral mucosa and to stimulate host immune responses through photobiomodulation of barrier epithelial tissue. Epithelial surfaces, including the oral and airway mucosa, are primary portals of entry for viruses and serve as the first line of host defense during infection. The interleukin-1 (IL-1) family of cytokines produced in epithelial tissue in response to invading microorganisms are apical cytokines that signal multiple downstream processes to affect both innate and adaptive immunity. Specifically, increases in IL-1α and IL-1β can signal key innate immune functions including triggering neutrophil recruitment, driving emergency myelopoiesis, and altering epithelial barrier permeability.^22^ Additionally, IL-1α and IL-1β are biologically active at very low concentrations and have previously been demonstrated to play a critical role in the control of influenza virus-related disease.^23^

Following photobiomodulation of uninfected oral epithelial tissues with 425 nm blue light (Figure 2C), the preferential upregulation of IL-1 cytokines and simultaneous decline of IL-6 and IL-8 response (IL-6, IL-8) is a significant finding, particularly due to IL-6’s association with the development of severe COVID-19.^24^ Transcription of inflammatory cytokines is known to be upregulated following oxidative stress induced by UV light through NF-κB pathways.^25^ However, the absence of changes in oxidative and cellular stress response biomarkers and the lack of any increase in IL-6 and IL-8 classically expressed as part of NF-κB transcription, suggest that 425 nm visible light is activating transcription pathways contrary and independent from the crosstalk of reactive oxygen species and NF-κB signaling reported for UVB (280-320 nm) and UVA (320-400 nm) wavelengths.^15,25^

Importantly, as a light-based treatment regimen, photobiomodulation of IL-1α and IL-1β can occur before tissue resident immune cells can recognize virus, potentially stimulating the target tissue to a more highly infection resistant state before virions are subsequently introduced to respiratory epithelial tissues. Kagan and co-workers have provided support for the natural function of IL-1 cytokines beyond their fundamental role in innate immune signaling.^26^ Mechanistic studies in epithelial cells revealed IL-1 cytokines can induce an antiviral state in surrounding tissue through promotion of interferon stimulated genes (ISGs) as a backup antiviral mechanism during encounters with immune-evasive viruses. The ineffective antiviral interferon (IFN) responses mounted by the immune system following SARS-CoV-2 infection has been recognized as a signature of COVID-19 disease pathology and contributes to aggressive disease progression.^27^ As such, safe photobiomodulation with 425 nm light of the IL-1 cytokines provides a host defense mechanism against SARS-CoV-2 that can prime other cells in advance of infection and via a pathway that is insensitive to immune evasion strategies used to prevent IFN gene expression.

In the early feasibility clinical trial, use of the investigational RD-X19 device in the outpatient setting resulted in a reduction in SARS-CoV-2 saliva viral load and a corresponding reduction in time to COVID-19 disease resolution. As summarized in Table 2, numerous outcome assessments resulted in a treatment benefit following 4 days of twice daily dosing with the RD-X19. The -1.48 log_10_ greater reduction in saliva viral load for RD-X19 compared to sham at Day 8 is clinically meaningful (Figure 4A), for both the primary subject as well as those potentially impacted by viral shedding and secondary transmission.

As suggested by FDA guidance for the development of therapeutics for COVID-19, a post-hoc analysis was conducted on symptom scores using a “sustained resolution of symptoms” definition.^19^ The 57-hour reduction in median time to sustained symptom resolution shown in Figure 5B demonstrates the robust clinical benefit from the RD-X19 treatment regimen. For comparison purposes, if confirmed in a subsequent trial with a larger sample size, this level of treatment benefit exceeds the clinical benefit demonstrated for approved drugs to treat otherwise healthy subjects with acute uncomplicated influenza (e.g. baloxavir marboxil, oseltamivir phosphate).^28,29^ The reduction of SARS-COV-2 viral load at day 8 and the large proportion of study participants with sustained resolution of all COVID-19 illness (85% for RD-X19 compared to 55% for sham) in this study demonstrate that a reduction of viral load in the saliva can lead to a clinically meaningful improvement in subject outcomes.

While the overall mortality rate for COVID-19 has declined in the United States due to better care for hospitalized patients, the number of global cases propagated from variants of concern continues to climb. Nearly a year ago, officials within the National Institutes of Health concluded that the availability of therapeutics as a complement to vaccines, especially those administered easily at home and widely available, ‘would have significant implications for the ability to end this pandemic.^30^ Monoclonal antibody cocktails have been granted Emergency Use Authorization that allow healthcare providers to administer these therapies to non-hospitalized patients with confirmed COVID-19 who are experiencing mild to moderate symptoms and are at high-risk for severe symptoms and hospitalization. Clinical trials for oral antivirals for at-home use are ongoing (e.g. molnupiravir), but they too are targeting the high risk population. Currently, there are no FDA authorized/approved treatment options available for the 75% of all outpatients diagnosed with COVID-19 that have no underlying risk factors.^9^ The potential advantages of RD-X19 for outpatient treatment include its safety, ease of administration in an at-home setting, and variant-agnostic mechanism of action.

The study described herein was originally designed as a hypothesis generating, proof-of-concept study with a small sample size. However, this early feasibility study evaluating RD-X19 as an early intervention (≤ 3 days from symptom onset) for adult outpatients with COVID-19 was successful in establishing proof-of-concept for a safe, at-home dosing schedule of 16 J/cm^2^ twice daily (128 J/cm^2^ total dose). Future trials will aim to further evaluate the safety and efficacy of RD-X19 with a larger sample size and may examine other clinical outcomes of cardiopulmonary function in targeted patient populations.

## Data Availability

All publicly available data produced in the present work are contained in the manuscript.

## Acknowledgements

We thank the trial participants; their families; the investigational site members involved in this trial; the members of the safety monitoring committee [Evyan Cord-Cruz, M.D. (Symbio, LLC), G Stephen DeCherney, M.D., M.P.H. (UNC Health), Jay Stevens, M.D. (Essential Health)], program management (Beth Landis and Jeff Hunter), Mike Bergmann, PhD for contributions toward the optical modeling of RD-X19 in the oral cavity, Michael Lay for graphic support, and Cari Green for her assistance with the assembling the manuscript.

## Author Contributions

Drs. Stasko and McNeil had full access to all of the data for the study and take responsibility for the integrity of the data and the accuracy of the data analysis.

*Wrote Manuscript:* Stasko, McNeil

*Designed Research:* Stasko, Cockrell, Emerson, McNeil

*Performed Research/Data Acquisition:* Jones, Santander, Henderson, Wood, Bradrick, Kocher, Henson

*Data analysis and interpretation:* Wang, Smith, Stasko, Cockrell, Emerson, McNeil

