## Supplementary Information for "A randomized, controlled, feasibility study of RD-X19 in patients with mild-to-moderate COVID-19 in the outpatient setting"

**EB-P12-01 Study Sites, Investigators, and Additional Study Personnel**

**EB-P12-01 Inclusion and Exclusion Criteria**

**Supplemental Methods**

**Supplemental Figures**

**Supplemental Tables**

This appendix has been provided by the authors to give readers additional information about their work.

**EB-P12-01 Study Sites, Investigators, and Additional Study Personnel**

APF Research, LLC (Miami, FL)

PI: Jorge L Santander. M.D., Ph.D.
Sub-I: Ramon F. Sastre, M.D.
CRC: Felix Perez, FMD BSN
CRC: Pedro J. Marrero, FMD BSN
CRC: Yaima Ortega, FMD
CRC: Mariana Girod
CRC: Tania Gonzalez
CRC: Anabel M. Iznaga

J&S Studies, Inc. (College Station, TX)

PI: Terry Jones, M.D.
Clinical Director: Jeremy Scott
Carla Clanton, RN
CRC: Lindsey Horton
CRC: Brent Clary
CRC: Melissa Moye

MRI Global (Kansas City, MO)

Ryan Howard
Carolyn Culp
Robert Krumsick
Theresa Seitz
Joy Miller

**EB-P12-01 Inclusion and Exclusion Criteria**

A subject must meet all the following criteria to be eligible for **inclusion** in this study:

1. Positive for SARS-CoV-2 antigen via nasal swab at or within the past 24 hours of the screening visit as detected using an FDA authorized SARS-CoV-2 antigen test.
2. Onset of signs and symptoms consistent with COVID-19* no longer than within the past 3 days and have either a) fever of at least 100 °F or b) at least two moderate or severe symptoms (cough, sore throat, nasal congestion, headache, chills/sweats, muscle or joint pain, fatigue, and nausea) at the time of screening.
3. Provides written informed consent prior to initiation of any study procedures.
4. Be able to understand and agrees to comply with planned study procedures and be available for all study visits.
5. Agrees to the collection of nasopharyngeal swabs, oropharyngeal swabs, oral saliva specimen collection and venous blood specimens per protocol.
6. Agrees to refrain from using oral antiseptics (e.g. hydrogen peroxide rinse, Listerine) or mouthwashes of any kind during the study.
7. Male or non-pregnant female, 18 to 65 years of age, inclusive, at time of enrollment.
8. No uncontrolled disease process (chronic or acute), other than COVID-19 signs and symptoms*.
9. No physical or mental conditions or attributes at the time of screening, which in the opinion of the PI, will prevent full adherence to, and completion of, the protocol.

*The following COVID-19 onset of signs and symptoms generally appear 2-7 days after exposure to SARS-CoV-2:

- Cough
- Sore throat
- Nasal Congestion
- Headache
- Chills/sweats
- Muscle or joint pain
- Fatigue
- Nausea

Severity of each COVID-19 symptom will be assessed based on definitions used for graded adverse events:

- None (Grade 0): Not present
- Mild (Grade 1): Events that are usually transient and may require only minimal or no treatment or therapeutic intervention and generally do not interfere with the subject's usual activities of daily living.
- Moderate (Grade 2): Events that are usually alleviated with additional specific therapeutic intervention. The event interferes with usual activities of daily living causing discomfort but poses no significant or permanent risk of harm to the research subject.
- Severe (Grade 3): Events interrupt usual activities of daily living, or significantly affects clinical status, or may require intensive therapeutic intervention. Severe events are usually incapacitating.

A subject who meets any of the following criteria will be **excluded** from participation in this study:

1. Positive urine pregnancy test at screening.
2. Any medical disease or condition that, in the opinion of the site Principal Investigator (PI) or appropriate sub-investigator, precludes study participation.
3. Presence of self-reported or medically documented uncontrolled significant medical or psychiatric condition(s) other than COVID-19.
4. Reports a recent positive test result (within the past 6 months) for hepatitis B surface antigen, hepatitis C virus antibody, or HIV-1 antibodies at screening.
5. Has a history of alcohol abuse or other recreational drug (excluding cannabis) use within 1 month of Study Day 1.
6. COVID-19 signs associated with acute respiratory distress or imminent serious medical outcomes.^^
7. BMI ≥36.
8. Subjects living (e.g., siblings, spouses, relatives, roommates) in the same household cannot be enrolled.
9. Has participated in another investigational study involving any intervention for SARS-CoV-2/COVID- 19 within the past 6 months or any clinical trial with interventional investigational product within 30 days of screening.
10. Currently enrolled in or plans to participate in another clinical trial with an interventional investigational agent that will be received during the study period.
11. History of hospitalization within the past 60 days.
12. History of systemic antiviral therapies within the past 30 days.

13. History of oral corticoid steroid use within the past 14 days or steroid injection within the past 6 months. Active use of nasal or inhalable steroids is also exclusionary. Topical steroids are not exclusionary.
14. Has a history of hypersensitivity or severe allergic reaction (e.g., anaphylaxis, generalized urticaria, angioedema, other significant reaction) to nitrites, nitrates or sun exposure.
15. Has any oral abnormality (e.g. ulcer, oral mucositis, gingivitis) that in the opinion of the investigator would interfere with device use, or intra-oral metal body piercings that cannot be removed for the duration of the study. Metal orthodontia is permitted as braces will be covered by the device mouthpiece.

^^Potential Study Subjects Presenting with any of the following should be referred for immediate medical care and are not eligible for the study

- Fever > 104º F
- Cough with sputum production
- Rales and/or rhonchi
- Difficulty breathing or respiratory distress defined by a respiratory rate ≥30 per minute, heart rate ≥125 per minute, $SpO_2$ ≤93% on room air at sea level or $PaO_2/FiO_2$ <300.
- Persistent pain or pressure in the chest
- Confusion

### Supplemental Methods

#### *Method S1. Optical Modeling for Coverage of the Oropharynx and Surrounding Tissues with Light*

As shown in Figure 1A, the RD-X19 device delivers a dose (also referred to as fluence and reported in J/cm$^2$) of visible light to the oropharynx and surrounding tissues. The approximate spread of the light beam and coverage relative to anatomical features is indicated by the blue shading in the figure. The beam is shaped and projected onto the surfaces of the oral cavity by the optical elements included in the RD-X19 device. The various elements of the optical system have been optimized in an iterative fashion to provide a safe, nominally designed dose of 16 J/cm$^2$ to the oropharynx as well as safe doses of light to other regions of the oral cavity. The qualitative shape of the projection of the shaped beam onto the irregular features in the oral cavity has been explored through a combination of first-order optical modeling and empirical measurements. Here we describe the process used to construct the optical model for a single subject. Results of the modeling are presented in Figure 1B, and the modeling process is described below.

The first step in building the optical model was obtaining suitable physical models for the RD-X19 device and the oral cavity. Since the RD-X19 prototype devices used in the trial were manufactured from engineering drawings, obtaining the suitable physical model for the RD-X19 was straight-forward. The device model used to manufacture the prototype trial devices was simply imported into Blender, a computer-aided design program that is used for creating 3D models, from SolidWorks, a solid modeling computer-aided design and computer-aided engineering program. For the oral cavity, a three-dimensional computed-tomography (3D CT) scan from a 42-year-old male, designated patient C3N-02693, was obtained from The Cancer

Imaging Archive, a database of publicly available medical images which is maintained by the National Cancer Institute. This patient was chosen due to his age (42 years old vs. median age in the study of 40), the high resolution and contrast of the scan, the presence of the teeth (particularly the incisors) which are used in positioning and targeting the RD-X19, the lack of any medical devices implanted in the oral cavity that may have interfered with the scan, and the absence of cancer impinging on the oropharyngeal and nasopharyngeal regions. This scan was first segmented into anatomical sections for easier manipulation using a program called 3D Slicer, which is used for medical image analysis and visualization. The segmented scan was then converted to a 3D Computer-Aided-Design (CAD) model for the oral cavity in Blender. While the model of the oral cavity was simplified by excluding several anatomical elements (e.g. tongue), it did include all of the elements needed for positioning the device (e.g. upper jaw, lower mandible, and teeth) and for assessing the incident light dose on all relevant tissues (e.g. posterior oropharyngeal wall, inner cheeks, uvula, soft palate, tonsils, and epiglottis). Only elements that did not affect the optical simulation were removed. For example, since the tongue is shielded from the light beam by elements of the RD-X19 when the device is positioned in the oral cavity, its removal did not affect the simulation.

Next, the RD-X19 was positioned in the oral cavity. First, the lower mandible was opened as it would be when the RD-X19 is inserted into the oral cavity for clinical use. The device was subsequently located in the oral cavity using the positioning elements integrated into the RD-X19. These elements use the teeth, and particularly the incisors, to fix the device position and orientation relative to the posterior oropharynx. After the geometry of the combined model was mapped in Blender using Standard Triangle (or Tessellation) Language (STL), the combined model was decimated to reduce the tessellation count to a size that would be manageable for use in the subsequent optical simulations while preserving appropriate resolution in accordance with the size and morphology of the features to be resolved. A global coordinate system within this combined assembly was then created to provide a reference for the optical simulation.

The decimated and combined oral cavity/RD-X19 3D CAD model was then imported from Blender into LightTools, a Monte-Carlo-based ray-tracing program used for 3D simulations of complex illumination systems with the ability to incorporate volumetric optical effects, scattering, and absorption. Subsequently, each mesh tessellation was resolved into a number of integrating elements, or bins, to sum source ray intercepts. Each bin was assigned a color corresponding to the number of source ray intercepts in that bin, and bin size was chosen to produce a smooth and continuous colormap. Similarly, the number of source rays was chosen so that the number of samples collected by each bin would be statistically significant. For example, the oropharynx was divided into one-hundred sixteen-thousand bins and included nearly three-million ray intercepts. Before the optical simulation was run, optical properties such as absorption and scattering coefficients were assigned to the device and anatomical surfaces as appropriate for the materials out of which the RD-X19 is constructed and for the mucosal membranes present in the oral cavity.

To connect the optical simulation obtained with LightTools with real-world RD-X19 device application, the optical simulation was anchored to the light dose nominally designed to be delivered to the oropharynx (16 $J/cm^2$). Namely, the maximum light dose at the tessellation representing the oropharynx in the combined model was scaled to 16 $J/cm^2$. The factor required to make this adjustment was applied to the entire 3D optical map of the oral cavity. As noted, a contour map of the light dose applied to the oral cavity during a single treatment cycle with the RD-X19 is shown in Figure 1B.

Simulation results were then compared to emission from a representative RD-X19 device at five anatomical regions of interest: oropharynx, uvula, epiglottis, tonsil, and soft palate. For this comparison, the coordinate system used in the model was replicated in the laboratory setting, and light values were measured at specified coordinates in three-dimensional space on the laboratory bench. Empirical measurements were performed using a Thorlabs S142C Integrating Sphere Photodiode Sensor with a 1 mm diameter pinhole attached to the aperture. In the comparison of simulated results to bench top measurements, we found that that model correctly identified the anatomical regions of highest (oropharynx, epiglottis, and uvula) and lowest (tonsil and soft palate) dose correctly. Further, the average error between the simulation and the bench measurements was found to be 7.2%, well within expectations given the complexity of the optical model and the difference.

As shown in optical simulation in Figure 1B, the RD-X19 device supplies a relatively smooth and uniform dose of light without 'hot spots' to the challenging and complex geometry of the oropharynx and surrounding tissues. Of note, the modeled maximum light dose supplied to any tissue in the oral cavity was found on the epiglottis to be roughly 18.7 $J/cm^2$ (measured at 19.5 $J/cm^2$ in the bench measurement), substantially lower than the minimal erythemal dose of 82 $J/cm^2$ at 425 nm.[2] While the work described here represents a single RD-X19 used in a single subject, we have further generalized the modeling presented to include variation in devices and the anatomical variation of the oral cavity found among adults.[1] In this work, we also found that the maximum dose supplied by the RD-X19 at any location in the oral cavity was below the minimal erythemal dose, regardless of the size of the oral cavity studied. This finding was substantiated by the results of the trial in which the dose of light supplied by the RD-X19 was found to be safe and well-tolerated across the range of subjects enrolled.

1. Urbizu A, Ferre A, Poca MA et al. (2017). Cephalometric oropharynx and oral cavity analysis in Chiari malformation Type I: a retrospective case-control study. *J Neurosurg*. 2017;126(2): 626–633. DOI 10.3171/2016.1.jns151590
2. Moseley H, Naasan H, Dawe R, Woods J, Ferguson J (2009). Population reference intervals for minimal erythemal doses in monochromator phototesting. *Photodermatol. Photoimmunol. Photomed.*, 25(1): 8-11, DOI: 10.1111/j.1600-0781.2009.00391

***Method S2. RT-PCR Determination of SARS-CoV-2 Viral Load***

SARS-CoV-2 RNA was quantitated using RT-qPCR on total RNA extracted from subject saliva samples. Saliva was collected from each subject at the clinical site in the FDA, EUA approved OM-505 (DNA Genotek) device. Samples were stored at room temperature prior to shipping to MRIGlobal for subsequent extraction and analysis. Upon receipt at MRIGlobal the samples were pre-processed prior to extraction by adding 0.5 mL of Qiagen PowerBeads (cat. #60941-064) to the sample and bead beating for 2 minutes at max speed using a Vortex-Genie 2. Bead beating of saliva samples ensures homogeneity prior to RNA extraction. RNA was extracted following manufacturer's instructions for the QIAamp Viral RNA Mini Kit (spin protocol) with a single modification (elution with 140 uL of buffer). Individual aliquots of RNA were then stored at -80 °C prior to quantification by RT-qPCR.

Quantification of SARS-CoV-2 RNA was performed using the N1 primer/probe set as defined by the 2019-nCoV CDC EUA Kit procured from Integrated DNA Technologies and ThermoFisher TaqPath 1-step RT-qPCR master mix. Each sample was concurrently evaluated bythe RNase P assay as an endogenous control to confer the presence of RNA isolated from each participants saliva sample. Accordingly, primer-probes for this assay were also procured in the 2019-nCoV CDC EUA kit as a control. All samples for the N1 and RNase P assays were evaluated in triplicate and collected as a cycle threshold (Ct) value. The N1 Ct values for each sample were quantitated against a standard curve prepared with synthetic SARS-CoV-2 RNA procured from ATCC (ATCC VR-3276SD).

*Quantitation of RT-PCR Results*

Each participant's sample was calculated as the concentration of RNA in genome equivalents (or copies) per mL. Samples with undetectable Ct values were assigned a value of 1. Only participants with detectable Day 1 (baseline) Ct values were included in the modified analysis population. All data were log transformed (log10) and assessed as change relative to baseline values. Normalized delta Ct values (ΔCt) were calculated from the difference between RNase P and N1 primer Ct values.

### Supplemental Figures

***Figure S1: Symptom Questionnaire* -** Representative single diary entry for self-assessment of COVID-19 signs and symptoms

Subject Initials: **[ENTER HERE]**
Subject Number: **[ENTER HERE]**

Study Diary

Protocol: EB-P12-01
Cohort 1

**Day 2 - 1st Dose** **#VALUE!**

**Please grade each of your symptoms below or mark None.**

| **Symptom** | **None** | **Mild** (can tolerate easily) | **Moderate** (interferes with daily activities) | **Severe** (prevents normal daily activities) |
|---|---|---|---|---|
| Nausea | ☐ | ☐ | ☐ | ☐ |
| Fatigue | ☐ | ☐ | ☐ | ☐ |
| Muscle or Joint Pain | ☐ | ☐ | ☐ | ☐ |
| Headache | ☐ | ☐ | ☐ | ☐ |
| Chills and Sweats | ☐ | ☐ | ☐ | ☐ |
| Sore Throat | ☐ | ☐ | ☐ | ☐ |
| Nasal Congestion | ☐ | ☐ | ☐ | ☐ |
| Cough | ☐ | ☐ | ☐ | ☐ |

Oral Temperature: ____________ F / C (circle one)

**Time of 1st Dose:** ______________ ☐ **Dose Missed**

(If treatment is interrupted, please allow the device to time out and complete treatment within 30 minutes)

**Comment:** ____________________________________________

*Figure S2: RNA Expression of Cellular/Oxidative Stress Markers and Housekeeping Genes analyzed via QuantiGene.*

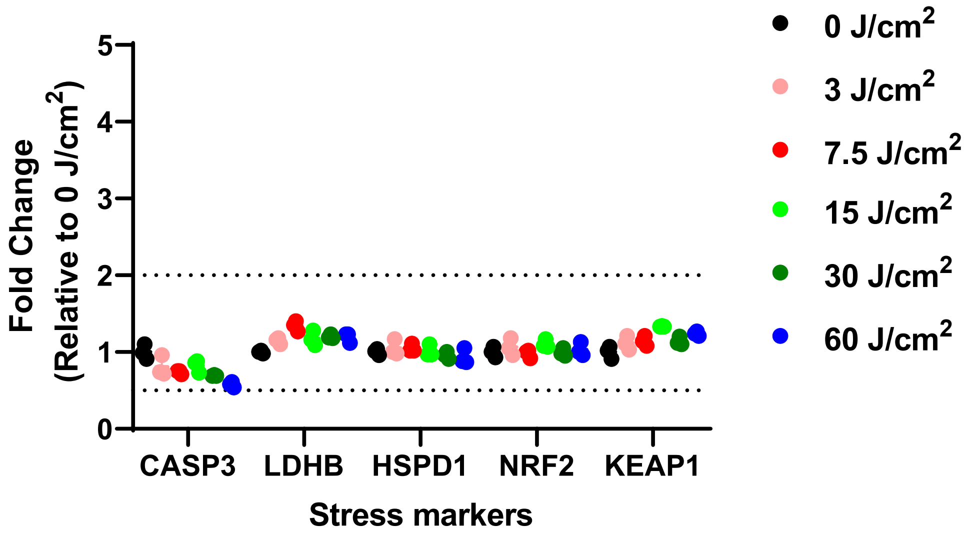


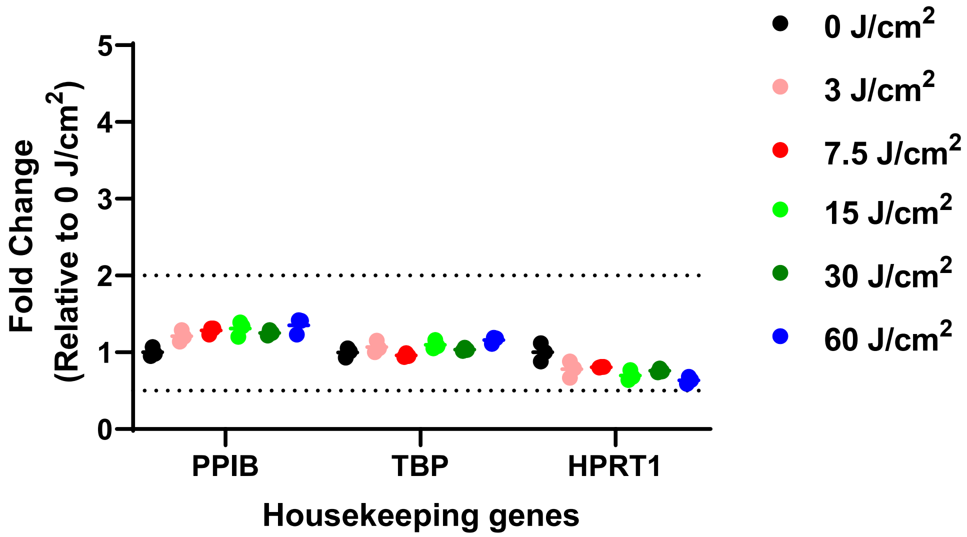

***Figure S3: Quantitative SARS-CoV-2 Copies/mL***

$Log_{10}$ transformed viral load (copies/mL +/- SEM) for all subjects at each visit in the modified analysis set having a positive saliva SARS-CoV-2 sample at baseline as detected via the N1 primer probe set via RT-qPCR (RD-X19 – blue circles and Sham – red squares).

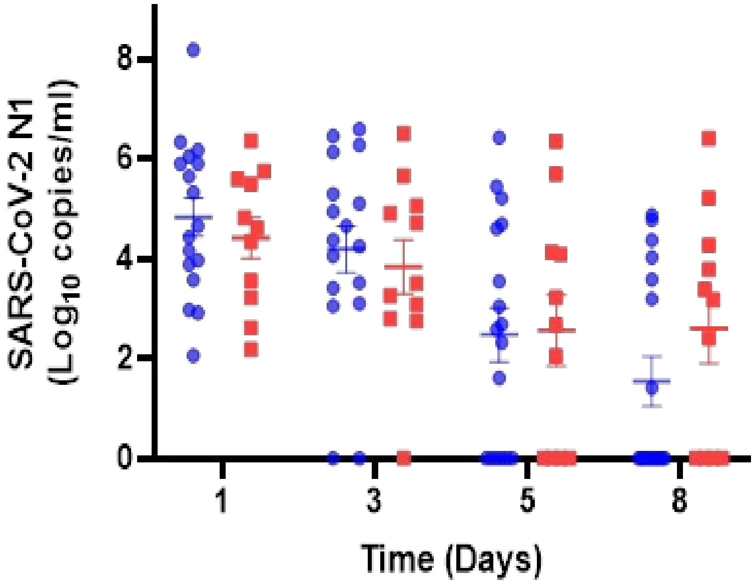

### Supplemental Tables

***Table S1. Summary of Treatment Emergent Adverse Events in the trial population.***

| | RD-X19 N=20 | SHAM N=11 |
|---|---|---|
| Number of participants, (percent) | | |
| **Serious Adverse Events** | **0 (0.0%)** | **0 (0.0%)** |
| **Severity of treatment emergent adverse events[a]** | | |
| Mild | 6 (30.0%) | 2 (18.2%) |
| Moderate | 3 (15.0%) | 4 (36.4%) |
| Severe | 3 (15.0%) | 3 (27.3%) |
| **Adverse events according to preferred term[b,c]** | | |
| Nausea | 5 (25.0%) | 3 (27.3%) |
| Chills | 0 (0.0%) | 3 (27.3%) |
| Fatigue | 2 (10.0%) | 2 (18.2%) |
| Myalgia | 3 (15.0%) | 0 (0.0%) |
| Headache | 1 (5.0%) | 1 (9.1%) |
| Cough | 0 (0.0%) | 1 (9.1%) |
| Nasal Congestion | 5 (25.0%) | 2 (18.2%) |
| Oropharyngeal Pain | 2 (10.0%) | 1 (9.1%) |

a. Includes the full randomized population that were dispensed a treatment device. A treatment emergent adverse event was defined as an event that first occurred or worsened in severity after baseline.

b. Any worsening or new signs and symptoms consistent with COVID-19 that first occur on or after study day 4 were recorded on participant self-assessment daily diary cards, documented as associated with COVID-19 by study subject, and attributed to COVID-19 disease pathogenesis by site investigators.

c. The preferred terms were defined according to the *Medical Dictionary for Regulatory Activities*, version 23.
